# Serious Workplace Violence against healthcare providers in China in the past 15 years

**DOI:** 10.1101/2020.05.20.20105072

**Authors:** Jing Ma, Xi Chen, Qiongjuan Zheng, Yun Zhang, Zhi Ming, Dongxin Wang, Hua Wu, Haisen Ye, Xiaoxuan Zhou, Yunxuan Xu, Renjiao Li, Xia Sheng, Fangxiu Fan, Zuiwen Yang, Ting Luo, Yajun Lu, Ye Deng, Fen Yang, Chuntao Liu, Chunyu Liu, Xiaosong Li

## Abstract

**Introduction:** Workplace violence (WPV) against healthcare providers has severe consequences and underreported worldwide. The aim of this study was to present the features, causes, and outcomes of serious WPV against healthcare providers in China.

**Method:** We searched serious WPV events reported online and collected information about time, location, people involved, methods used, motivations, and outcomes related to the incident, and analyzed their summary statistics.

**Result:** Serious WPV reported online (n=379) in China were mainly physical (97%) and often involved the use of weapons (34.5%). Doctors were victims in most instances (81.1%). WPV mostly happened in cities (90.2%), teaching hospitals (87.4%), and tertiary hospitals (67.9%), frequently in ED, OB-GYN, and pediatrics, in the months of June, May, and February. WPV Rates increased dramatically in 2014 and decreased after 2015. Death (12.8%), severe injury (6%), and hospitalization (24.2%) were the major outcomes.

**Conclusion:** Serious WPV in China may stem from poor patient-doctor relationships, overstressed health providers in the highly demanded hospitals, poorly educated/informed patients, insufficient legal protection and poor communications. A law protecting healthcare providers implemented in 2015 may have helped curb the violence.

## Introduction

Workplace violence (WPV) refers to an individual’s or group’s socially unacceptable, aggressive (and sometimes destructive) behavior.WPV against healthcare workers is a global public health problem that has been underreported and largely ignored[1].

WHO estimated that 8-38% of healthcare workers suffer from physical violence while working[2]. Many more are threatened or exposed to verbal aggression[3]. The damage due to workplace violence translates into physical and mental harm to the health workers. The research literature shows that such violence leads to issues such as death [4], heart and brain disease[5], anxiety, depression [6], and PTSD[7,8]. Workplace violence also translates to high costs for the organization where it takes place, both in the short- and long-term. It decreases the quality of care provided to all patients, not only violent ones. In China, workplace violence in hospitals causes a lot of medical students to change their majors and decreases the integrity of the healthcare provider-patient relationship [9].

The perpetrators who carry out violent behavior against healthcare workers vary with respect to their relationship to the worker: some are patients, some are patients’ relatives, while others are neither [10].Research literature from Greece and Nepal has shown nurses are more likely to be victims of WPV than doctors[11,12]and that verbal violence is more common than physical violence [3,13]. However, a study in China showed doctors are more frequently the victims [14]. Additionally, physical violence against doctors appears to be more common than physical violence against nurses in China[15]. There are only a few studies on WPV in China[11,13,16];the prevalence of WPV varies from province to province [15,17], from hospital to hospital[13,18], and from department to department[19,20]. China is the only country in which prevalence of WPV by month has been studied; according to previous research, it is most common in July[14].

Many researchers have tried to determine the reasons behind WPV, which can vary as a result of different medical systems and national conditions. There is a lot of literature that explores the outcomes of WPV[21,22]. Of all the countries with research on the topic, we found WPV in China leads to the most serious outcomes, such as death[4].

Serious WPV against healthcare workers, though less common than milder forms of violence, possibly gets more attention from mass media and the public. It shows the worst relationship between healthcare provider and patient, also reflects particularly negative living situations of healthcare providers in certain medical systems. It reveals the suffering and helplessness of patients, as well as the defects of certain medical and legal systems. Serious WPV usually happens suddenly, leading to high levels of conflict and dispersion, which makes research on the topic hard to carry out. Studying mass media reporting may therefore be a good way to study serious WPV.

As far as we know, there are have only been two studies about serious WPV against healthcare workers in China. There are a few more studies focusing on WPV that is not serious[4,14]. One of these articles shows the changes of prevalence and features of serious WPV against doctors and nurses in China, as reported online from 2000 to 2015 [4]. This article will present the newest changes in, features of, reasons for, and outcomes of WPV trends against healthcare providers in China from 2004 through 2018 based on online reports.

## Methods

The research data examined in this article came from online reports about workplace violence against healthcare workers in hospitals from January 2004 to December 2018. Baidu, Sogou, Souhu, and Lilac Garden were used as search engines, and “ShangYi” (do harm to doctors), “Yi Yuan” and “Bao Li”(hospital and violence), “Yi Nao”(medical harassment), “Da Yi Sheng”(beating doctors), “Da Hu Shi”(beating nurses), “Yi Huan Chong Tu”(healthcare provider-patient disputes), and “Bao Li Shang Yi”(healthcare workers’ injury by workplace violence) were used as search words for finding news and reports online.

All the data were came from online reports, this study did not involve any human subjects, animals or plants, so there is unnecessary for the Ethics approval and consent to participate.

Relevant online information was screened, and secondary materials were excluded. We read the reports and collected the following information about the violence: causes; time(year, month); region(province, city, county, town); hospital (name, public/private, level of the hospital if public); department; types of violence(verbal, physical, or both); identity of victims(doctor, nurse, other staff member); identity of perpetrators (patient, relative of the patient, other person); and outcomes of the events(death, injury, name of the injury, admitted to IPD or not). We asked a coroner to read the outcome information we collected and to determine how serious the injuries were (severe injury, minor wound, minor bodily injury).

SPSS17.0 was used to input data and to do statistical analysis. We used the frequencies command to evaluate frequency and proportion of WPV with regard to location(province, city, county, town, hospital, and department); time(top month and yearly changes); outcomes; reasons (losing control of emotions, dissatisfaction and high expectations for therapeutic outcome, unreasonable request for procedures); features of violent behavior; and identity of perpetrators and victims.

## Results

### Sample size

There were 379 violent events reported. Some information was not mentioned in the reports, which led to missing values. However, there was complete information on sample sizes of province, year, and name of hospital. The number of reports that included information for the remaining fields are as follows: department: 219, month:378, day:371, city: 368, hospital level: 258, teaching hospital or not: 364, identity of victim: 370, types of violence: 370, reasons: 372, with weapon or not: 365, identity of perpetrator:331, outcomes: 265.

Identity of victims and perpetrators and features of violent behavior

Doctors were among the victims in 300 events (81.1%), and nurses were among the victims in134 of the cases (36.2%), meaning that both types of providers were injured in 64 events (17.3%). There were 30 events in which other persons (security guards, policemen, etc.) were injured, too.

Relatives of the patients were the perpetrators in 190 events (57.4%), and patients themselves were perpetrators in 132 events (39.9%). Sometimes patients and their relatives carried out violent behavior together (12, 3.6%). There were 28 acts of violence (8.4%) committed by non-relatives of patients.

The reported violent events included physical violence (beating, slapping of the face, stabbing with knife, hitting with bricks/chairs, forcing victims to kneel, kidnapping, stalking, etc.) 97% of the time (n=359).Verbal violence (insulting, cursing, swearing, shouting, threatening, intimidating, etc.) occurred in 21.1% of the events (n=78). Almost one-fifth (n=67, 18.1%) of the events included both physical and verbal violence, and 34.5% of perpetrators used a weapon (knife, brick, stick, table, stairs, etc.).

### Location: province, city, county, town, hospital, department

There are 31 provinces and 4 municipalities in China, and none of them was free from workplace violence in the past 15 years, though the frequency of incidence varied from province to province. The five provinces/municipalities with the most WPV and the percentage of the total incidents that occurred in each are as followings: Guangdong: 52(13.7%), Hunan: 30(7.9%), Jiangsu: 25(6.6%), Beijing: 22(5.8%), Guangxi: 21(5.5%).

Most of the workplace violence in hospitals happened in cities(332, 90.2%), while only 36 events(9.8%) happened in counties and towns. Most of the workplace violence happened in tertiary hospitals (243, 67.9%), which are the highest ranking hospitals in the system, and only rarely in first-level hospitals (10, 2.8%)or private hospitals (8,2.2%). The remainder either happened in secondary hospitals (72,20.1%) or other public hospitals whose level was not mentioned(25,7.0%). Remarkably, of all the hospitals that reported violent workplace incidents, teaching hospitals accounted for 87.4% (318) while non-teaching hospitals accounted for 12.6% (46).

The top three departments with the highest rates of WPV were emergency departments (74, 33.8%), obstetrics-gynecology departments (26, 11.9%), and pediatrics (20, 9.1%). In total, 18.3% of events happened in a department related to internal medicine (n=40). A total of20.1% of events happened in departments related to surgery not associated with obstetrics-gynecology (n=44).

### Time: year, month

From 2004 to 2013, the incidence rate fluctuated. The incidence increased significantly over the year during 2014 and peaked in 2015, then decreased gradually in the following years. By the end of the timeframe of interest, rates had decreased to the lowest levels in the past five years, which were almost as low as the rates in 2013(See Fig1:WPV in the past 15 years).

**Fig 1:**
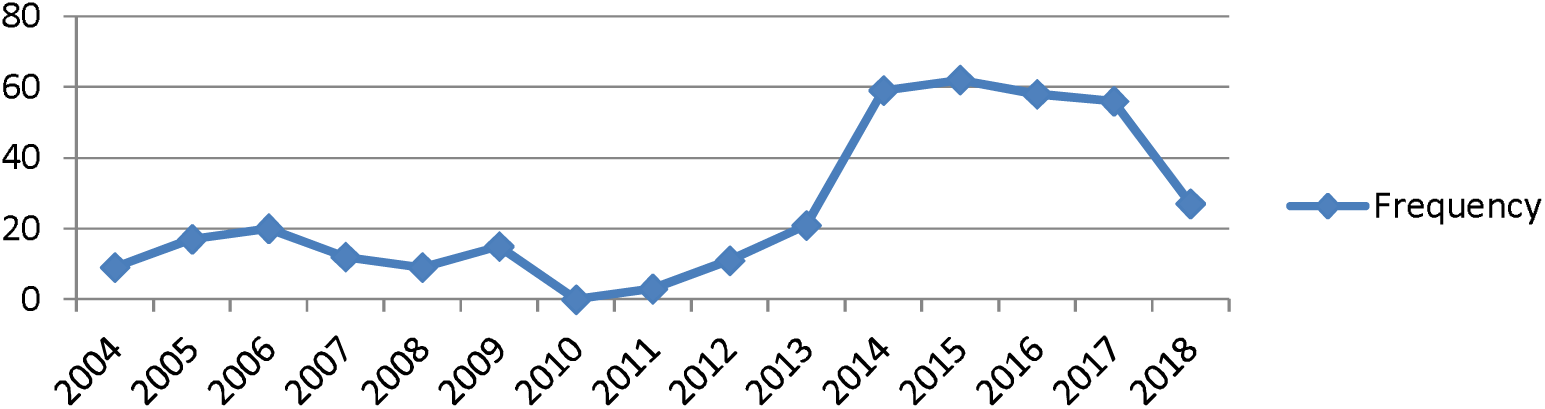
WPV in the past 15 years.

The five months with the most occurrences were: June (72, 19%), May (40, 10.6 July (35, 9.3%), and February (35, 9.3%)(See Fig2: Month distribution of WPV in the past 15 years).

**Fig 2:**
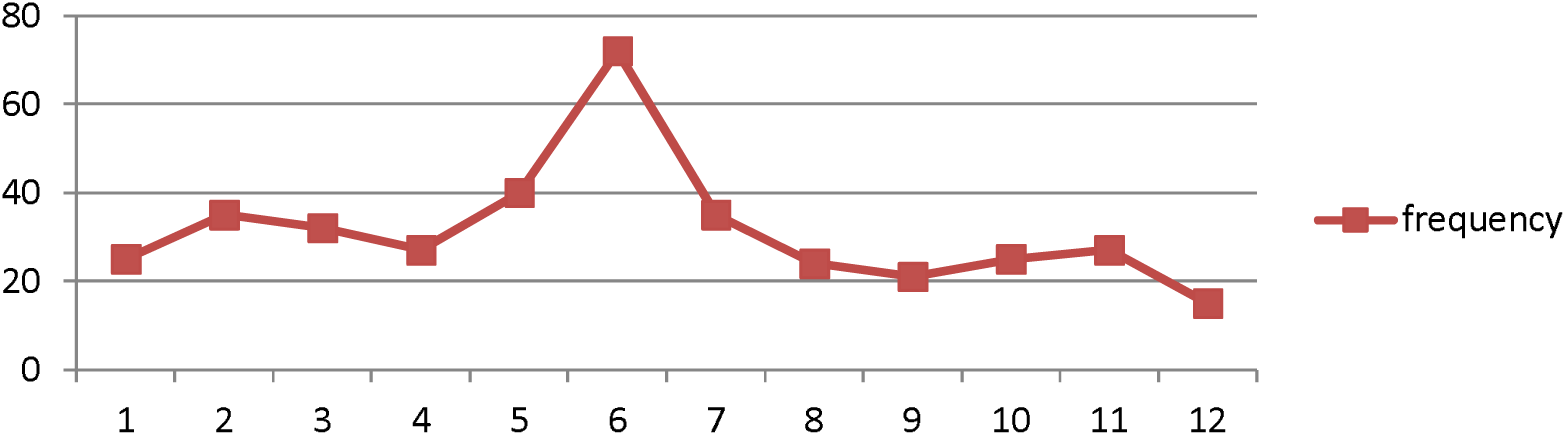
Month distribution of WPV in the past 15 years.

### Outcome of violent events

265 reports (69.9% of total sample) included information about the outcome of the injured persons. 171 out of 265 had detailed descriptions about injury severity and target body parts.

The severity of injuries was as follows: death(34, 12.8%); severe injury(16,6.0%) such as pierced heart, paralysis of both lower limbs, decapitated arm, or intestinal perforation; minor wound(70,26.4%) such as intracranial hemorrhage, orbital fracture, concussion, miscarriage, second-degree/third-degree burn, tendon rupture, or lung contusion; slight bodily injury(82,30.9%) such as light closed encephalon injury, threatened miscarriage, soft tissue contusion, nose bleeding, head trauma, facial blood stasis, or waist injury.

Moreover, the injured persons suffered head and face injuries in 102events (38.5%),trunk injuries in 33 events (12.4%), and limb injuries accounted for 6.1% (16) of injuries; notably, many people suffered multiple injuries in one event; 64 events(24.2%)led to hospitalization.

### Reasons for violent behavior

The reasons behind the perpetrators’ acts of violence in the hospital are displayed in the following table:

**Table 1:** Reasons for violent behavior

| Reasons | n | Proportion |
| --- | --- | --- |
| Reasons are not mentioned | 75 | 20.20% |
| Refusing to accept the death of the patient | 47 | 12.60% |
| Being not satisfied with the therapeutic effect | 39 | 10.50% |
| Thinking the emergency treatment is not effective | 27 | 7.30% |
| Wanting to get treatment as soon as possible without following medical procedures | 24 | 6.50% |
| Being drunk | 13 | 3.50% |
| Having suspected mental disorder | 12 | 3.20% |
| Believing that severe treating consequence is due to clinical operations | 11 | 3.00% |
| Failure clinical operation(such as puncture) of operating for the second time | 10 | 2.70% |
| Asking the professional staffs to give treatment and arrangement as they requested, but being refused | 40 | 1.80% |
| Being diagnosed as mental disorder | 5 | 1.30% |

## Discussion

The purpose of this study was to reveal the features of serious WPV reported online against healthcare deliverers in China. Our results convey insights into the people involved in, the times of, the locations of, as well as the methods used for and the outcomes of serious WPV in Chinese hospitals.

Who & How: Our study found the vast majority of cases of serious WPV reported online were physical in nature (97%) and were often committed with weapons (34.5%). Such a high occurrence of weapon use suggests extreme conflict between patients and healthcare providers. Our study also found that doctors were more exposed to serious WPV than nurses. Several previous studies showed nurses were more exposed to verbal violence than doctors[3,11,23,24]and that doctors were more often the victims of physical workplace violence[25,26]. These results indicate that the most serious WPV may result from more major issues such as those related to diagnosis and treatment—which are primarily linked to doctors—rather than from smaller issues related to nurse-patient interactions. Perpetrators are more often relatives of patients than they are patients themselves. This finding has been reported cross-culturally[27,28]. This may be because patients are sometimes unable to move or to argue or fight due to medical conditions and age. Relatives may express themselves through violence as a result of anger, worry, dissatisfaction, or financial intentions (claim for compensation).

Where: Guangdong, Hunan, Jiangsu, Beijing, and Guangxi had the highest rates of WPV. Qinghai, Hainan, Ningxia, Neimeng, Taiwan, and Shanxi had the lowest prevalence rates. Previous similar research in China reached similar conclusions, with the greatest prevalence occurring in Guangdong, Jiangsu, Sichuan, Zhejiang and the lowest rates occurring in Gansu, Ningxia, Tianjin, Shanxi, and Taiwan[4].We searched the populations and GDPs of the above provinces on National Bureau of Statistics of China (http://data.stats.gov.cn/search.htm) and found that the top provinces in WPV incidents have high GDPs or large populations. Most provinces that were amongst the lowest prevalence-wise had amongst the lowest GDPs or smallest populations in China. We speculate that economically developed provinces attracting millions of migrant workers every year—and thus adding to the already overloaded burden of the health providers by local residents—has led to a higher frequency of WPV. Such a great need for medical attention may strain medical staff resources and thus result in worse patient-doctor relationships, contributing to the prevalence of WPV. The relationship between these socioeconomic variables and WPV is complicated and requires further research.

This study found that serious WPV mostly happens in cities (90.2%) and usually occurs in tertiary hospitals (67.9%), especially teaching hospitals, which account for 87.4% of WPV events in tertiary hospitals. The finding regarding teaching hospitals differs from Chen’s conclusions, which showed the incidence of WPV in teaching hospitals was lower than the incidence of WPV in regional hospitals in China and was similar to incidence in developed countries[29]. Difference in methods and regions of interest may account for the discrepancy between these findings. Some studies [4,15,30]such as Yan’s report on Heilongjiang have shown that Chinese tertiary hospitals usually have higher rates of WPV than hospitals in rural areas or small towns[30]. The current study found similar results. It is worth mentioning that tertiary hospitals in cities of China usually are best equipped also with the best doctors, where patients with comparatively severe, challenging diseases usually seek help. That means that at these kinds of hospitals, the death toll per year can be expected to be higher, increasing motivation behind WPV. Furthermore, almost every doctor in teaching hospitals faces a great pressure to do research and publish articles in order to get a promotion, which forces them to reduce clinical hours. Routine service in the inpatient units of teaching hospitals is mostly performed by resident trainees, postgraduate students, and further educational doctors. These doctors have less experience interacting with patients; fewer medical skills and abilities, which may raise their likelihood of getting into medical disputes[31].

According to our findings, the three departments making-up the highest proportion of WPV incidents were ED, OB-GYN, and pediatrics. Emergency departments have been previously described as being at high risk for violent incidents[4,14,19,26,27,32], a finding that was corroborated by the current study. Samir found 86.1% of nurses in OB-GYN departments had been exposed to WPV[28]. Li Z found that Chinese healthcare providers in children’s hospitals experienced violence commonly and that 68.6% of staff members had experienced at least one WPV incident in the past year[27]. Ferri found that the top three departments for WPV were psychiatry (86%), emergency(71%), and geriatric wards (57%).[24]However, Min’s study from China showed that the frequency of WPV in OB-GYN (9) and pediatrics (7) was not higher than in other internal medicine and surgical departments [4]. We think that the high number of incidents reported in OB-GYN departments and pediatrics may have to do with the dramatic increase in maternal and child care hospitals (primarily pediatrics and OB-GYN), as well as children’s hospitals in China in the past five years.WPV occurring in these Children-related hospitals increase the counts in those related departments.

When: We found serious WPV increased dramatically in 2014 and decreased gradually after 2015. This shift may be the result of an article added to the Criminal Law of the People’s Republic of China (ninth revision) in 2015. The article reads: “Where people are gathered to disturb public order to such a serious extent that work in general, production, business operation, teaching or scientific research cannot go on and heavy losses are caused, the ringleaders shall be sentenced to fixed-term imprisonment of not less than three years but not more than seven years; the active participants shall be sentenced to fixed-term imprisonment of not more than three years, criminal detention, public surveillance or deprivation of political rights.”

The three months during which WPV occurred most frequently in the past 15 years were June, May, and February (Table 1). February is usually the month in which Spring Festival occurs. During Spring Festival, there is a shortage of staff members in hospitals, which may heighten risk of medical disputes. A report published in China by Yuqing found that the top three months for WPV were May, June, and July[14]. No research thus far has revealed the reasons behind the inordinately high amounts of WPV in June and May. This may be a good area of research for future study.

Outcome: WPV has very serious consequences. We are shocked by the rate of death (12.8%), severe injury (6%), and hospitalization (24.2%) that has resulted from WPV. Previous research in developed countries has reported that WPV has more frequently resulted in non-physical harm[5-8,33]. On the contrary, research in China--including this study--has found that physical harm is more common; these instances of physical violence have sometimes led to death[4]. We are strongly worried about the safety of doctors in China. It is worth mentioning the injuries that were classified as “minor” were categorized according to a forensic standard in China and that many of these injuries--such as abortion, renal contusion, coma, and hemorrhagic shock--would not be considered minor by most people.

Reasons: We separated the reasons we found for WPV into three categories:

1. Losing control of emotions, including:”being drunk”, “having a diagnosed or suspected mental disorder”. Previously, Bataille found alcohol abuse is one of the most common triggers of WPV in ED[34]; a lot of other research has similarly found drunkenness and mental disorders are often associated with physical violence against healthcare providers[24,35-38].
2. Dissatisfaction and high expectations for therapeutic outcome, including: “failed clinical operation(like puncture)”, “operating for the second time”, “thinking the emergency treatment is not effective”, “believing that severe treatment consequence is due to clinical operations”, “refusing to accept the death of the patient”, and “being dissatisfied with the therapeutic effect”. The dissatisfaction of therapeutic outcome was due to two reasons: either actual poor quality of medical care or unreasonable expectations leading to dissatisfaction in the face of normal medical failures or flaws. Previous research showed similar results pertaining to ineffective treatment and high expectations related to WPV[39], but the level of physical harm we reported was more serious. We speculate that these intense conflicts in China stem, in part, from negative healthcare provider-patient relationships and a lack of relevant legal measures. Both of these issues may be consequences of flaws in the medical system. This also may be a good area of research for future study.
3. Unreasonable requests for procedures, including: “asking the professional staff to give treatment and arrangements as requested but being refused” and “wanting to get emergency treatment as soon as possible without following medical procedures.” Alkorashy found misunderstandings and long waits for service are factors that contribute to WPV.[40]Inadequate professional resources and poor communication between healthcare providers and patients may also sometimes be the reason behind unreasonable request for procedures[26].

## Limitation

The main limitation of this study was that it was based on online reports, whose integrity and authenticity was influenced by factors such as government regulations, areas where reports were made, the interests of public media and Internet companies, the professional ethics of the journalists responsible for the reports, and the validity of the resources. There is a chance that some incidents that occurred in rural areas and under-developed regions were not reported and thus not included in this study, which could bias some of the analyses.

## Conclusion

The current findings reflect a bleak healthcare setting in China, dangerous conditions for healthcare workers, and poor doctor-patient relationships, which may, in large part, be due to problems with the Chinese medical system, including overstressed health providers in the highly demanded hospitals, poorly educated/informed patients, insufficient legal protection and communications between care-providers and patients. We strongly believe that public education should be improved to reduce patients’ unreasonable expectations. Furthermore, better allocation of medical resources and more legal action against WPV could reduce serious workplace violence.

## Data Availability

The data is available from the corresponding author.

**Abbreviations**
WPV: Workplace violence

## Acknowledgments

We would like to thank Tanya Horwitz for word revision.

